# Inequities in Effective Coverage of Family Planning Services in Low-and Middle-Income Countries: Linking Households and Facility Surveys

**DOI:** 10.1101/2025.02.13.25322079

**Authors:** Shenning Tian, Elizabeth A Hazel, Melinda Munos, Abdoulaye Maïga, Safia S Jiwani, Emily B Wilson, Gouda Roland Mesmer Mady, Agbessi Amouzou

**Affiliations:** Department of International Health, Johns Hopkins Bloomberg School of Public Health, Baltimore, MD USA

## Abstract

**Background:** Despite decades of family planning (FP) program successes in low-and middle-income countries (LIMCs), unmet need for contraceptives persists with inequalities in coverage. Including elements of service readiness in family planning intervention coverage measures will better inform population-level program performance.

**Methods:** Five LMICs countries were identified with a health facility and household survey conducted within five years of one another within the last 10 years: Bangladesh, Haiti, Malawi, Nepal and Tanzania, with time trend data available for Nepal. We used ecological linking methods to develop quality readiness adjusted FP coverage measures by linking health facility assessments (readiness) and household surveys (intervention coverage). Linking units were defined by facility type, managing authority and geographic location using women’s reported source of contraceptives. Intervention coverage was defined as the percentage of women 15-49 years of age in need of contraceptive services who were using a modern method and we used the average FP readiness score in each linking unit to calculate readiness-adjusted intervention coverage. We used a coverage cascade model to understand gaps in health service readiness and access, and performed a health equity analysis for wealth, locality and age.

**Findings:** We found large gaps in FP intervention coverage and readiness in all settings. Facility readiness scores ranged from 0.60 and 0.75 with gaps in coverage and readiness-adjusted coverage ranging from 49 percentage points (pp) in Bangladesh to 34pp in Malawi. Urban, wealthier and adolescent women had lower readiness-adjusted coverage since they also obtained their contraceptives outside of health facilities. We found little change in the coverage cascades for Nepal from 2015 to 2021.

**Conclusions:** This analysis demonstrates calculating readiness-adjusted FP coverage using a cascade model in five geographically diverse countries with time trends in one country. As previous studies have shown, we found large gaps in intervention and readiness-adjusted coverage with related inequalities.

## Background

Effective family planning (FP) programs improve maternal and child health by reducing high-risk or unwanted pregnancies, unsafe abortions, and promote socioeconomic development through reducing poverty, increasing women’s educational opportunities, and advancing human rights and gender equality. Despite substantial improvements in FP programs over the last 50 years, in 2021, 164 million women of reproductive age worldwide still had an unmet need for contraception. [3] Most women facing unmet needs are from low- and middle-income countries (LMICs), with more than half living in sub-Saharan Africa and South Asia.[4]

One important factor contributing to unmet need for family planning is the poor quality of care. A study has shown that in 2015, 27% of women in LMICs reported discontinuation of contraception due to poor quality of care.[5] Sufficient quality of FP services enables informed choice of contraceptives, appropriate knowledge of use and management of contraceptive side effects and continuity of care.[6] Using classic quality of care frameworks, quality encompasses structural, process and outcomes.[7] Structural quality – also called readiness – measures whether the environment of health services is sufficient to provide quality care. Process quality is the standard of care and outcomes are patient level results such as knowledge and satisfaction with services. The linkages between these three domains is not linear, likely due to the complex nature of health service provision. [8] This study focuses on measuring readiness (structural quality) as a necessary but insufficient condition of process quality and improved patient outcomes.

A common indicator of FP service coverage is the proportion of women using contraception among those who are in need of services does not reflect the quality of services delivered. [9] To address this limitation, effective coverage measures have been proposed to add quality of care elements to coverage to better capture the potential health benefits of the intervention. [10–12] Amouzou and colleagues proposed a coverage cascade framework consisting of six steps to measure the potential loss of health benefits at each step.[13] Using this stepwise model, we define the target population, service contact, intervention coverage and readiness-adjusted coverage for demand for family planning services satisfied from a health facility equipped and ready to provide quality FP services (table 1)

**Table 1.** Coverage Cascade and definitions

| Coverage Cascade | Definition |
| --- | --- |
| Services contact | Woman aged 15-49 who is currently using a modern method OR woman was met by a health worker at facility for other care or field worker who mentioned FP in previous 12 months |
| Intervention coverage | Woman aged 15-49 in need of contraceptives who is currently using a modern contraceptive from any source |
| Intervention coverage, at health facility | Woman aged 15-49 in need of contraceptives who is currently using a modern contraceptive sourced at a health facility (excludes private pharmacies, shops, etc.) |
| Readiness-adjusted coverage | Woman who is currently using modern contraceptives obtained at health facility equipped to provide contraceptive services |

The objective of our study was to describe the facility readiness and inequities in readiness-adjusted coverage for FP care in Bangladesh, Haiti, Malawi, Nepal, and Tanzania. In Nepal where multiple data were available, we also assessed trends in readiness and coverage cascade of FP services.

## Methods

### Data

This study was a secondary analysis of the publicly available data from the Service Provision Assessments (SPA) and the Demographic and Health Surveys (DHS) in Bangladesh, Haiti, Malawi, Nepal and Tanzania. The SPA is a sample survey or census of health facilities that collects national and sub-national level information on the availability and quality of health services from the country.[14–19] The DHS was conducted in a nationally representative sample of households to provide estimates on demographic and health indicators, including access and use of modern family planning methods, at the national and sub-national levels.[20–25] These five countries were selected because they had DHS and SPAs with the required data that were completed within five years of each other since 2015 (table 2). Nepal was the only country with two SPA and DHS that met this criteria to perform a trend analysis.

**Table 2.** Description of the surveys

|  |  |  |  |  | Contraceptives Currently Use (%) |  |  |  |  |  |  |  |  |
| --- | --- | --- | --- | --- | --- | --- | --- | --- | --- | --- | --- | --- | --- |
| Country | Survey Type | Survey Year | Sampling | Sample size | Pill | IUD | Injections | Male Condom | Sterilization | Implants/Norplant | Emergency Contraception | Traditional | Other Modern* |
| Bangladesh | SPA | 2017 | Sample | 1,353 |  |  |  |  |  |  |  |  |  |
|  | DHS | 2017-18 | Sample | 18,895 | 41.00 | 0.89 | 17.38 | 11.61 | 9.53 | 3.45 | <0.01 | 16.09 | <0.01 |
| Haiti | SPA | 2017-18 | Sample | 756 |  |  |  |  |  |  |  |  |  |
|  | DHS | 2016-17 | Sample | 14,371 | 5.82 | 0.25 | 49.95 | 25.92 | 3.28 | 5.64 | 0.00 | 7.59 | 1.50 |
| Malawi | SPA | 2013-14 | Census | 810 |  |  |  |  |  |  |  |  |  |
|  | DHS | 2015-16 | Sample | 24,562 | 3.78 | 1.75 | 48.96 | 5.59 | 18.27 | 19.49 | <0.01 | 1.75 | 0.16 |
| Nepal | SPA | 2015 | Sample | 899 |  |  |  |  |  |  |  |  |  |
|  | DHS | 2016 | Sample | 12,862 | 8.70 | 2.68 | 16.83 | 8.01 | 38.84 | 6.27 | 0.12 | 18.43 | <0.01 |
|  | SPA | 2021 | Sample | 1,478 |  |  |  |  |  |  |  |  |  |
|  | DHS | 2022 | Sample | 14,845 | 7.76 | 2.22 | 16.04 | 7.99 | 30.23 | 10.47 | <0.01 | 25.12 | <0.01 |
| Tanzania | SPA | 2014-15 | Sample | 933 |  |  |  |  |  |  |  |  |  |
|  | DHS | 2015-16 | Sample | 13,265 | 12.75 | 2.11 | 30.58 | 12.01 | 7.96 | 17.14 | <0.01 | 16.34 | 0.93 |

\*Includes lactational amenorrhea, female condom, standard days method, or other modern method

### Indicators

This study examined the readiness scores and coverage cascade of FP service. Services readiness scores were calculated from the SPA using FP tracer indicators identified according to the Service Availability and Readiness Assessment (SARA) guidelines, a tool aimed to assess and monitor the service availability and readiness of the health sector.[26] Based on the SARA, the tracer indicators were categorized into four domains: availability, staff and guidelines, equipment, and medicines and commodities (table 3). For Haiti, Nepal, and Tanzania, we removed the tracer indicator “progestin-only contraceptives” from the medicine and commodities domain because less than 30% of facilities in each country had them in stock, assuming these commodities were not a key part of the national program. Each domain was a binary or weighted average (0-1) with the total service readiness score a weighted average (table 3).

**Table 3.**
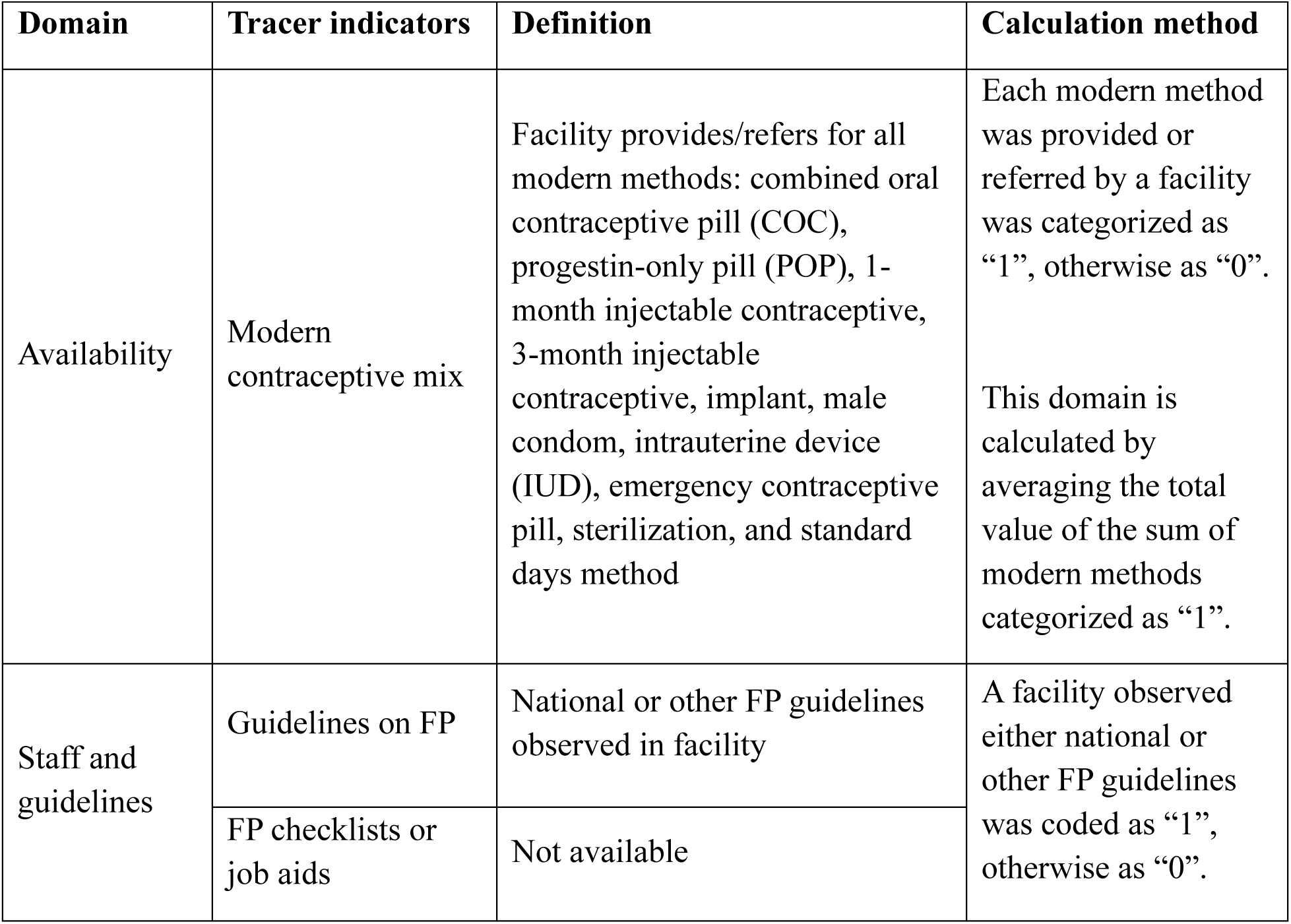
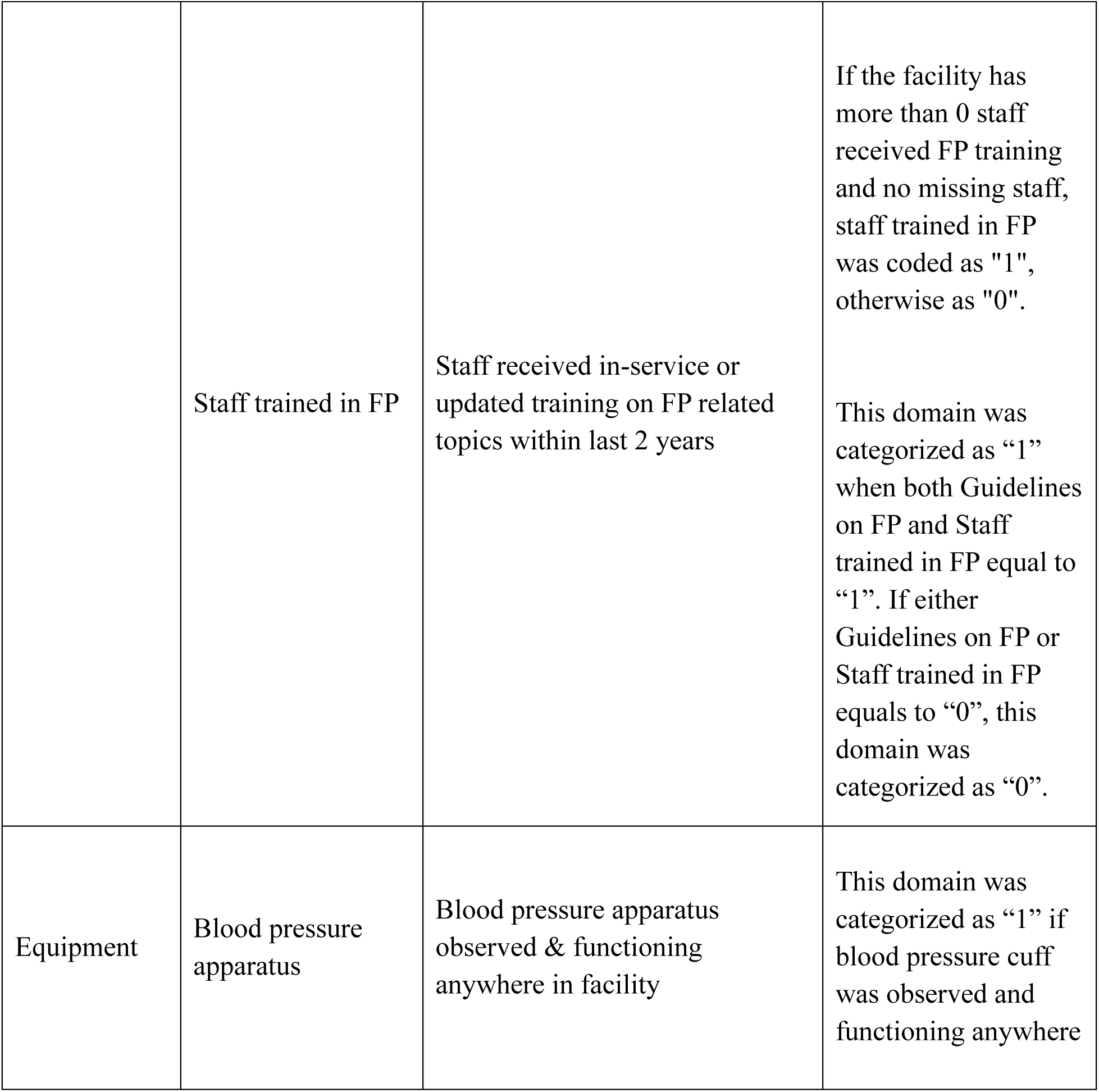
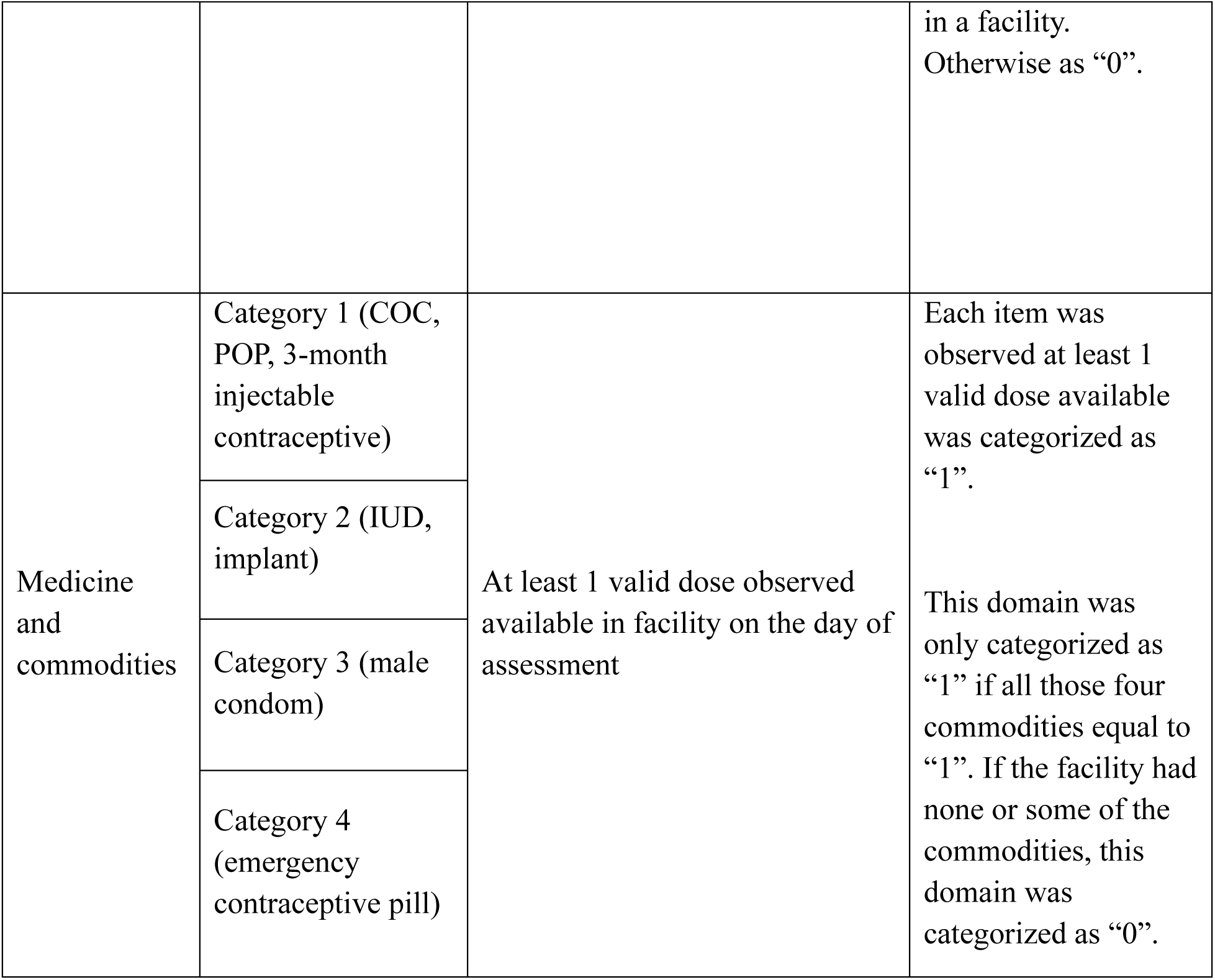
Domains and Their Tracer Indicators of FP Service Readiness

We applied the coverage cascade framework proposed by Amouzou et al in this analysis to quantify the use of FP services at different conditional stages. In this study, we focused on service contact (woman aged 15-49 who is currently using a modern method OR woman was met by a health worker at facility for other care or field worker who mentioned FP in previous 12 months), intervention coverage (demand for family planning satisfied with modern methods), intervention coverage at health facility (modern method was sourced at health facility), and readiness-adjusted coverage (table 1). Service contact, intervention coverage, and intervention coverage at health facility were directly calculated from the DHS data. Readiness-adjusted coverage is estimated by linking data from the DHS and SPA using the ecological linking methodology, which uses facility type (e.g. hospital or clinic), managing authority (public or private), and administrative area (region or urban/rural locality) as the unit to link data from the DHS and SPA. [27,28]

### Independent variables and data analysis

We categorized managing authority into public/government, private for-profit, and private non-for-profit (appendix 1). Public facility types were classified as hospital, health center, health post/clinic, and community health workers. Private for-profit facilities included private hospitals, private health centers, private health posts/clinics, and private community health workers. We categorized any type of non-governmental organization or mission/faith-based facility under private non-for-profit managing authority. Sources such as pharmacies, shops, and churches were not considered as formal sources and quality is set to zero. Community health workers were not included in these SPAs and this analysis does not account for them.

We categorized the geographical areas of Bangladesh, Haiti, and Tanzania into urban and rural since there were insufficient number of facilities per linking unit to generate a readiness score. Malawi was divided into northern, central, and southern regions, while Nepal’s geographic areas were categorized by ecological regions into mountain, hill, and terai. We performed a comparative analysis to determine disparities in FP service readiness and coverage by comparing the service readiness components and coverage cascade across the five countries. We then disaggregated the coverage cascade by geographical area (urban and rural), women’s age, and wealth quintile, and further analyzed the absolute and relative gaps between intervention and readiness-adjusted coverage for an equity analysis. The data were analyzed with STATA 18.0 (16).

## Results

The most common contraceptive method varies by country. Injectable contraceptives were the most prevalent in Haiti (50.0%), Malawi (49.0%), and Tanzania (30.6%). The pill was most commonly used in Bangladesh (41%). In Nepal, sterilization was the predominant method although decreasing in use (from 38.8% in 2015 to 30.2% in 2021) (table 2). The source for modern contraceptives differed across countries, but the public sector was the main source (appendix 2).

The national FP service readiness score ranged from 0.60 in Nepal to 0.74 in Tanzania (table 4). Bangladesh had the highest availability of modern methods (0.78) compared to the other four countries (ranging from 0.57 in Tanzania and 0.64 in Nepal 2015). (Table 4). All countries had high availability of blood pressure cuffs in the facility (>0.86 score). Tanzania had the highest score for FP staff or guidelines (0.85) while Nepal had the lowest (0.62 in 2015 and 0.34 in 2021). Tanzania had the highest stock all four categories of commodities (0.58) whereas Haiti had the lowest (0.29). In Nepal – the only country with time trend data – the overall service readiness score stayed the same (0.68 in 2015 and 0.60 in 2021) but components of the score changed. The stock of all four categories of commodities in the facilities decreased from 0.50 in 2015 to 0.43 in 2021, as did FP staff/guidelines score: 0.62 in 2015 to 0.34 in 2021.

**Table 4.**
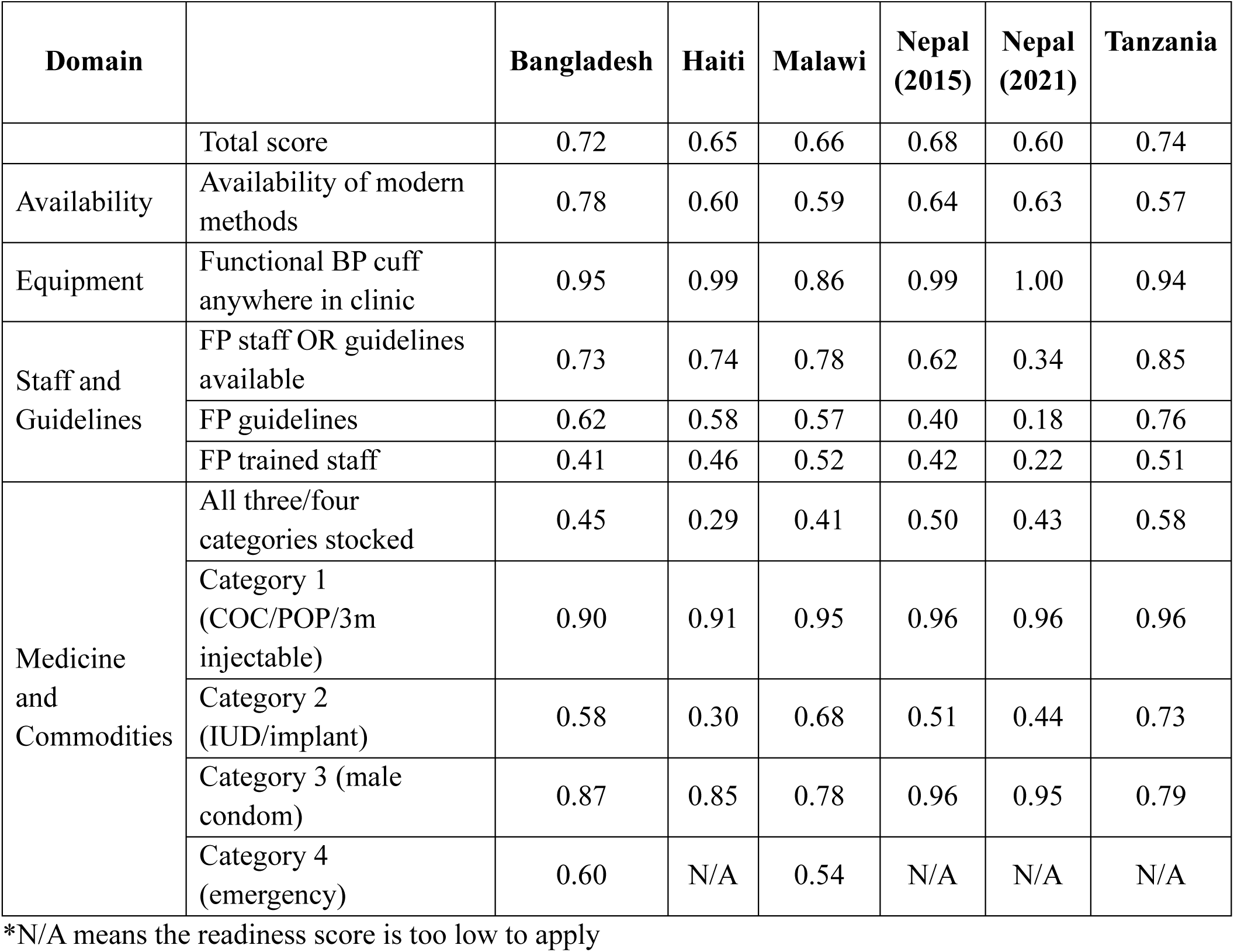
Services Readiness Component Scores

| Domain |  | Bangladesh | Haiti | Malawi | Nepal (2015) | Nepal (2021) | Tanzania |
| --- | --- | --- | --- | --- | --- | --- | --- |
|  | Total score | 0.72 | 0.65 | 0.66 | 0.68 | 0.60 | 0.74 |
| Availability | Availability of modern methods | 0.78 | 0.60 | 0.59 | 0.64 | 0.63 | 0.57 |
| Equipment | Functional BP cuff anywhere in clinic | 0.95 | 0.99 | 0.86 | 0.99 | 1.00 | 0.94 |
| Staff and Guidelines | FP staff OR guidelines available | 0.73 | 0.74 | 0.78 | 0.62 | 0.34 | 0.85 |
|  | FP guidelines | 0.62 | 0.58 | 0.57 | 0.40 | 0.18 | 0.76 |
|  | FP trained staff | 0.41 | 0.46 | 0.52 | 0.42 | 0.22 | 0.51 |
| Medicine and Commodities | All three/four categories stocked | 0.45 | 0.29 | 0.41 | 0.50 | 0.43 | 0.58 |
|  | Category 1 (COC/POP/3m injectable) | 0.90 | 0.91 | 0.95 | 0.96 | 0.96 | 0.96 |
|  | Category 2 (IUD/implant) | 0.58 | 0.30 | 0.68 | 0.51 | 0.44 | 0.73 |
|  | Category 3 (male condom) | 0.87 | 0.85 | 0.78 | 0.96 | 0.95 | 0.79 |
|  | Category 4 (emergency) | 0.60 | N/A | 0.54 | N/A | N/A | N/A |

\*N/A means the readiness score is too low to apply

Among the five countries, public facilities had the highest levels of readiness (appendix 3). However, the readiness levels of specific facility types varied by country and geographical area. In Haiti, Malawi, Nepal, and Tanzania, public hospitals had the highest readiness levels, ranging from 0.79 to 0.91, regardless of the geographical area. In Bangladesh, the readiness levels were highest in public *upazila* health complexes, with scores of 0.89 and 0.94.

Across the five countries, the percentage of women in need of FP services who had contact with FP services ranged from 60% in Haiti to 86% in Malawi (fig. 1). The proportion of women meeting their FP needs with modern contraceptives from any source varied from 45% in Haiti to 77% in Malawi. The percentage of women obtaining modern contraceptives from formal health sectors ranged from 34% in Haiti to 75% in Malawi. In all countries, the readiness-adjusted coverage was much lower: median 24.5pp difference between intervention and readiness-adjusted coverage in the five countries.

**Figure 1.**
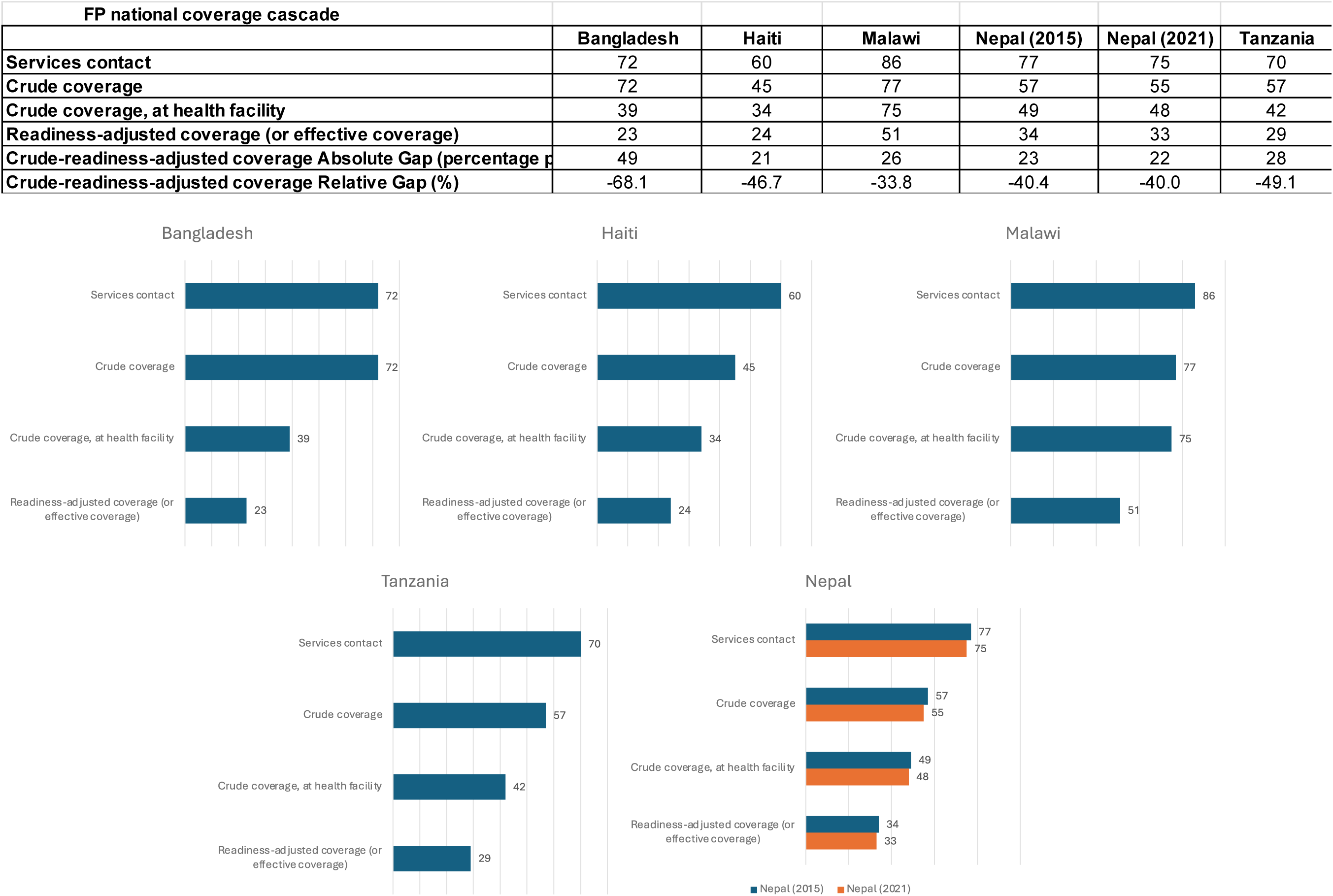
Family Planning National Coverage Cascade

We observed little variation across urban/rural locality in Bangladesh, Haiti and Tanzania or across geographic regions within Malawi and Nepal (fig. 2). In Bangladesh, Haiti, and Tanzania, intervention coverage was slightly higher in urban areas than rural areas, but readiness-adjusted coverage was lower in urban areas compared to rural areas. Many women in urban areas are obtaining their contraceptives outside the formal health sector in these urban areas. Bangladesh had the largest intervention-readiness-adjusted gaps in both urban (57pp, 76% relative reduction) and rural areas (45pp, 64% relative reduction). In Malawi, the absolute and relative gaps were similar across regions, ranging from 19 to 28pp, with reductions of 27% to 35%. Nepal showed more similar absolute and relative gaps, with little change between 2015 (21-24pp; 38%-43% reduction) and 2021 (22-23pp; 36%-45% reduction).

**Figure 2.**
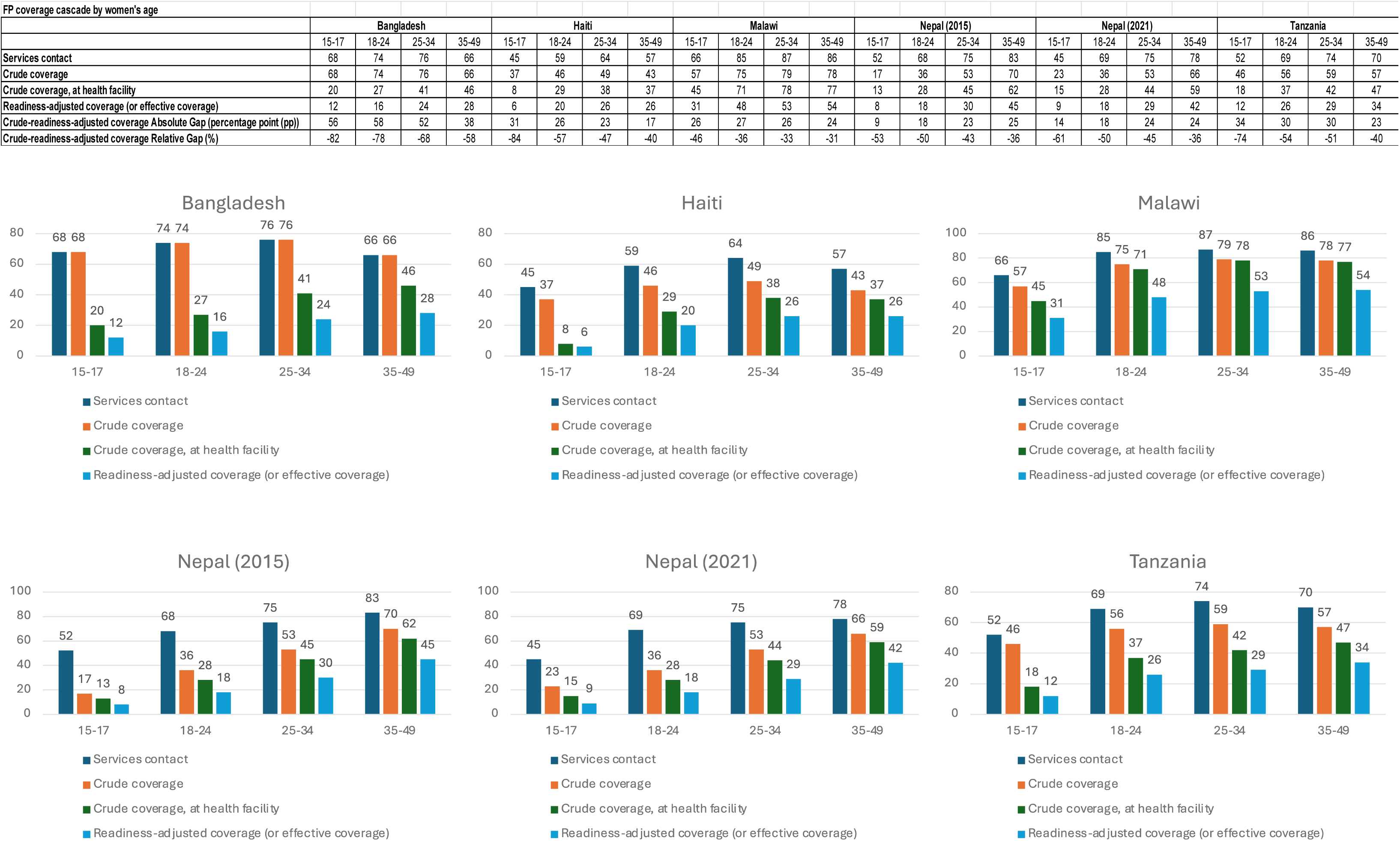
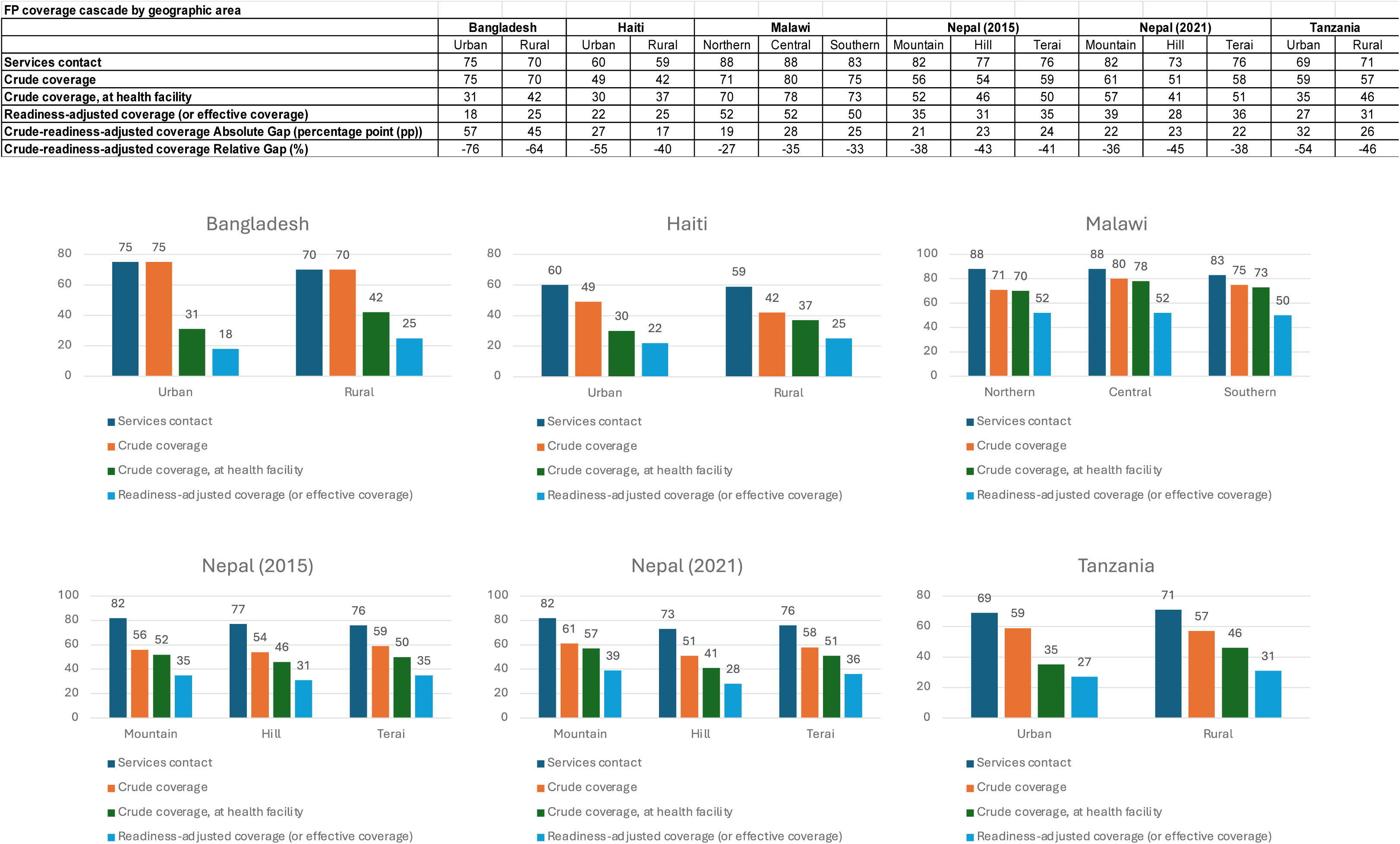
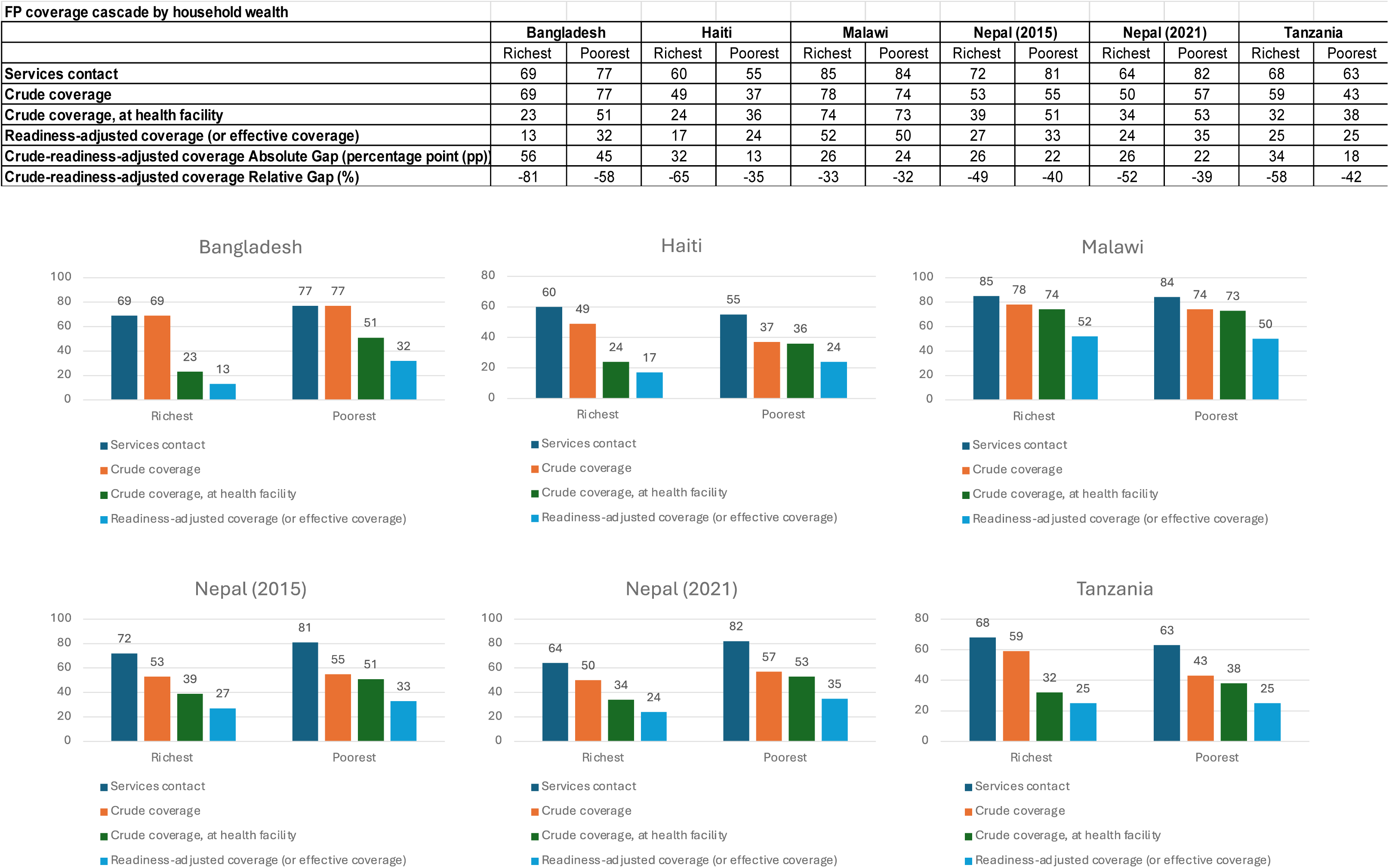
Family Planning Coverage Cascade by Geographic Area, Women’s age, and Household Wealth

Coverage at each stage of the cascade for adolescents (15-17 years of age) was lower than for the other three adult groups (fig. 2). The exception was Bangladesh, where service contact and intervention coverage for adolescents (68%) was similar for the 35-49 age group (66%). Intervention coverage for adolescents ranged from 17% in Nepal in 2015 to 68% in Bangladesh, and readiness-adjusted coverage ranged from 6% in Haiti to 31% in Malawi. Adolescents aged 15–17 had the largest relative gaps, with Haiti showing the largest reduction (84%), followed by Bangladesh (82% reduction), Tanzania (74% reduction), Nepal (61% /53% reduction), and Malawi (46% reduction). Most adolescent girls/women were obtaining their contraceptives from outside of health facilities.

We also found that the wealthiest women had higher intervention coverage in Haiti (49%), Malawi (78%), and Tanzania (59%) than the poorest women (37%, 74%, and 43%, respectively). However, in Bangladesh, the richest women (69%) had lower intervention coverage than the poorest women (77%). A similar pattern was observed in Nepal. When adjusting for facility service readiness, the richest women in Malawi and Tanzania had similar readiness-adjusted coverage than the poorest women, while in the other three countries, the wealthiest women had lower readiness-adjusted coverage than the poorest women, again due to wealthier women sourcing contraceptives outside of health facilities. Among the five countries, Bangladesh had the largest intervention-readiness-adjusted coverage gaps for the richest women (56pp; 81% reduction). In Nepal the gap is half of the intervention coverage and was maintained between 2015 and 2021.

## Discussion

This study analyzes facilities’ FP services readiness and readiness-adjusted coverage and equity in Bangladesh, Haiti, Malawi, Nepal, and Tanzania. Other studies have described family planning service readiness using these datasets but ours is the first to focus on the coverage cascade by linking health facility and household survey to describe the gaps and equity across a variety of settings.[29,30] We found that the overall score for facility readiness for FP services in the five countries ranged between 0.60 and 0.75 with large gaps between intervention coverage and readiness adjusted coverage. In some settings, the cascade shows the discrepancy in intervention and adjusted coverage is because women are obtaining contraceptives outside of health facilities (informal shops or community health workers not included in SPA surveys). This is seen particularly in Bangladesh where intervention coverage is 72% but only 39% obtained their contraceptive from a facility. Less so in Malawi, where the coverage-readiness gap is due more to low facility readiness. We also found urban, wealthier and adolescent women had lower readiness-adjusted coverage since they also obtained their contraceptives outside of health facilities. Finally, we were able to assess time trend in the coverage cascade for Nepal from 2015 to 2021 and found little change in the cascade but important differences in some readiness score domains (FP staff/guidelines score decreased from 0.62 in 2015 to 0.34 in 2021). Interpretation of time trend analysis scores should take into account actual changes in the national programs and questionnaire wording or differences in the survey protocol.

In this study, among all facility types, public hospitals had the highest readiness scores across the five countries, consistent with findings from previous studies. [29–31] This might be due to public hospitals receiving substantial investment from government sources and at the same time are required to comply with government policies to improve the readiness of FP services in order to provide quality services. Additionally, hospitals tend to be located in urban areas and more equipped and staffed than health centers.[32]

Among the five countries, Bangladesh had the largest coverage-readiness gap. Despite relatively high intervention coverage in Bangladesh, the large gap was driven by two factors. First, a substantial number of women sourced their contraceptive outside health facilities. Second, facilities offering FP services were not always ready. In Malawi, on the other hand, most contraceptive are sourced from health facilities, and therefore the gap between intervention coverage and readiness-adjusted coverage is due to low facility readiness for FP service. Studies have shown that women’s choice of obtaining FP services was often influenced by geographic distance and cost, with a preference for facilities that were closest to their residence and offer free contraceptives. [33–35]

Our study also assessed inequalities in coverage cascades by geographic region, women’s age, and household wealth. We found that women living in urban areas in Bangladesh, Haiti, and Tanzania had higher intervention coverage than women living in rural areas, which was consistent with findings from other studies. [36] However, readiness-adjusted coverage was lower for women living in urban areas than for those living in rural areas but the readiness scores of facilities in urban areas were not lower than those in rural areas Therefore, these differences may result from a large number of women living in urban areas relying on informal facilities for contraceptives (appendix 4). For instance, in Haiti, 37% of women in urban areas obtained their contraceptives from informal health facilities, compared to only 12% of women in rural areas.

Adolescents had the lowest levels of readiness-adjusted coverage with a linear increase by increasing age of the women (Fig. 2). Our study also revealed that adolescents were much more likely than adults to obtain modern contraceptives outside health facilities (appendix 4). Previous studies have shown that lack of privacy and confidentiality, as well as discriminatory and judgmental attitudes of healthcare providers, were major barriers that prevent adolescents from accessing contraception from health facilities.[37–40] Finally, we found that wealthier women had lower coverage along the cascade in Bangladesh and Nepal and lower readiness-adjusted coverage in Haiti. This may be due to FP programs targeting poor women in Bangladesh and Nepal, in Haiti, wealthier women are obtaining contraceptives outside the health facilities.

The major strength of this study is that we were able to link health facility assessments with household survey data to generate the coverage cascades in geographically diverse LMICs, including time trend in Nepal. Our assessment of the facility readiness and coverage cascade of FP services, as well as the stratifications of coverage cascade, can help identify weak domains of facility services and population groups with low intervention and readiness-adjusted coverage. This will help inform future interventions and policies regarding FP to improve health inequalities.

One of the limitations of our study is the temporal difference in when the DHS and the SPA were conducted. For example, the Malawi SPA data was from July 2013 to February 2014, whereas the Malawi DHS collected data between October 2015 and February 2016. The service readiness of facilities may change over time. Another limitation is the lack of consensus on the definitions and measurements of facility readiness. The SARA tracer items have been used in other studies but there is no demonstrated linkage between service readiness and FP outcomes. We used ecological linking method to link household and facility survey data, which linked a woman to an average readiness score but did not represent the readiness of the facility she actually visited. Some studies found that the ecological linking can produce similar outputs as direct linking.[41] Finally, these facility survey assessments did not include community-based health workers or private pharmacies. These health service delivery points may offer high levels of service readiness but since we did not have data for them, we are assuming a readiness of zero.

## Conclusion

Health facilities in Bangladesh, Haiti, Malawi, Nepal, and Tanzania showed moderate and variable facility readiness levels for providing FP services. This suggests that many women accessing health facilities for FP services might not have receive the quality of care expected. Improving readiness-adjusted coverage requires not only enhancing the readiness of health facilities, but also addressing barriers that hinder women, especially adolescents, from accessing modern contraceptives. Efforts should focus on promoting health equity to ensure that all women, regardless of socioeconomic status, have equal access to high-quality FP services.

## Ethics approval

This is secondary analysis of de-identified publicly available data. No one on the team has access to the personal identifiers linked to this data. Therefore, we do not consider this human subject research.

## Authorship contributions

The concept was originated by EAH and AA. ST conducted the analysis and wrote the first draft of the manuscript with review and input from all authors. MM, AM, SSJ, EBW and GRMM made important technical contributions to this analysis and are listed in order of the magnitude of their contributions. All authors approved the manuscript for publication.

## Data Availability

The data used in this study are publicly available from the Demographic and Health Surveys (DHS) Program at https://dhsprogram.com/Data/upon registration and approval.

## Acknowledgments

We would like to thank all the members of the Countdown to 2030 team at Johns Hopkins Bloomberg School of Public Health for their inputs including Assanatou Bamogo, Robert Black, Jennifer Requejo, Malick Kante, Shelley Walton, and Emma Williams. We also acknowledge and thank the participants who responded to the facility and household questionnaires used in this study.

