## Appendix 1 for "Inequities in Effective Coverage of Family Planning Services in Low-and Middle-Income Countries: Linking Households and Facility Surveys"

**Appendix 1. Provider Crosswalk**

| Country | Category | Provider type | Survey 1 | Survey 2 |
|---|---|---|---|---|
| Bangladesh | | | SPA 2017 | DHS 2017-18 |
| | Public | Government/public hopistal | district hospital | medical college hospital |
| | | | | specialized govt hospital |
| | | | | district hospital |
| | | Government/public UHC | upazila health complex (UHC) | upazila health complex |
| | | Government/public MCWC | Mother and Child Welfare Center (MCWC) | Mother and Child Welfare Center (MCWC) |
| | | Government/public UnHFWC | union health and family welfare center (UnHFWC) | union health and family welfare center |
| | | | union health and family welfare center (UnHFWC - Upgraded) | |
| | | | union subcenter (UnSC) / rural dispensary | |
| | | Government/public health clinic/post | community clinic | community clinic |
| | | | | satellite clinic/epi outreach |
| | | | | other public sector |
| | | | | government field worker (fwa) |
| | Private | Private for profit | private hospital | private medical college hospital |
| | | | | private hospital |
| | | | | private clinic |
| | | | | qualified doctor's chamber |
| | | | | non-qualified doctor's chamber |
| | | Private for non-profit (local govt/NGO) | ngo clinic (other than smiling sun clinics) | ngo static clinic |
| | | | ngo hospital | ngo satellite clinic |
| | | | smiling sun clinic of ngo health service delivery project (NHSDP) | ngo depo holder |
| | | | | ngo field worker |
| | | | | other ngo sector |
| | Not a health faciliy | Not a health faciliy | | pharmacy/drug store |
| | | | | shop |
| | | | | friend/relative |
| | | | | other |
| | | | | don't know |
| Haiti | | | SPA 2017-18 | DHS 2016-17 |
| | Public | Gorvenment/public hospital | university hospital | government hospital |
| | | | department hospital | mix hospital/clinic |
| | | | community reference hospital | |
| | | | other hospitals | |
| | | Gorvenment/public health center | health center with lit | government health center |
| | | | health center without lit | mix health center |
| | | Gorvenment/public health clinic/post | dispensary/community health center | mix family planning clinic |
| | Private | Private for profit | university hospital | private hospital, clinic |
| | | | community reference hospital | private health center |
| | | | other hospitals | family planning clinic |
| | | | health center with lit | private doctor |
| | | | health center without lit | |
| | | | dispensary/community health center | |
| | | Private for non-profit | department hospital | mobile clinic (ngo) |
| | | | other hospitals | health worker (ngo) |
| | | | health center with lit | |
| | | | health center without lit | |
| | | | dispensary/community health center | |
| | Not a health facility | Not a health facility | | pharmacy |
| | | | | shop |
| | | | | slot-machine |
| | | | | firend/relative |
| | | | | other |
| Malawi | | | SPA 2013-14 | DHS 2015-16 |
| | Public | Government/public hopistal | central hospital | government hospital |
| | | | district hospital | |
| | | | government/public rural/community hospital | |
| | | | government/public other hospital | |
| | | Government/public health center | government/public health center | government health center |
| | | Government/public health clinic/post | government/public maternity | government health post/outreach |
| | | | government/public dispensary | mobile clinic |
| | | | government/public clinic | hsa |
| | | | government/public health post | cbda/door to door |
| | Private | Private for profit | private other hospital | private hospital/clinic |
| | | | private health centre | private mobile clinic |
| | | | private clinic | prvate cbda/door to door |
| | | | private maternity | private doctor |
| | | | private dispensary | |
| | | | company health centre | |
| | | | company dispensary | |
| | | | company clinic | |
| | | Private for non-profit | cham rural/community hospital | cham/mission hospital |
| | | | cham other hospital | cham/mission health center |
| | | | cham health centre | cham/mission mobile clinic |
| | | | cham maternity | cham/clinic door to door |
| | | | cham dispensary | blm |
| | | | cham clinic | macro |
| | | | mission/faith-based other hospital | youth drop in center |
| | | | mission/faith-based clinic | |
| | | | ngo health centre | |
| | | | ngo clinic | |
| | Not a health faciliy | Not a health faciliy | | other private medical |
| | | | | pharmacy |
| | | | | shop |
| | | | | church |
| | | | | friend/relative |
| | | | | other |
| | | | | don't know |
| Nepal | | | SPA 2015 | DHS 2016 |
| | Public | Gorvenment/public hospital | central government hospital | government hospital/clinic |
| | | | regional government hospital | |
| | | | sub-reginal government hospital | |
| | | | zonal government hospital | |
| | | | district government hospital | |
| | | | other public hopital | |
| | | | central level government hospital | |
| | | | district level government hospital | |
| | | Gorvenment/public health center | primary health care center | primary health care center |
| | | | urban health centre | |
| | | Gorvenment/public health clinic/post | health post | health post/sub health post |
| | | | sub-health post | primary health care outreach clinic |
| | | | | mobile camp |
| | | | | female community health volunteer |
| | | | | other public facilities |
| | | | | institutionalized fp clinics |
| | Private | Private for profit | other hospital | private hospital/nuring home |
| | | | | private clinic |
| | | | | sangini outlet |
| | | Private for non-profit | other hospital | family planning association of nepal |
| | | | htc | marie stopes |
| | | | | other ngo facilities |
| | Not a health facility | Not a health facility | | other private medical facilities |
| | | | | pharmacy |
| | | | | shop |
| | | | | firend/relative |
| | | | | other |
| | | | | don't know |
| | | | SPA 2021 | DHS 2022 |
| | Public | Gorvenment/public hospital | federal level hospital | government hospital |
| | | | provincial level hospital | phc/primary hospital |
| | | | local level hospital | |
| | | Gorvenment/public health center | primary health care center (phcc) | basic health care center |
| | | | community health unit (chu) | community health unit |
| | | | urban health centre (uhc) | urban health center |
| | | Gorvenment/public health clinic/post | health post (hp) | health post |
| | | | | mobile camp |
| | | | | female community health volunteer (fchv) |
| | | | | other public sector |
| | | | | institutionalized fp clinics |
| | Private | Private for profit | other hospital | private hospital |
| | | | | private clinic |
| | | | | sangini outlet |
| | | Private for non-profit, mission/faith-based | other hospital | family planning association of nepal (fpan) |
| | | | htc | marie stopes |
| | | | | other ngo medical sector |
| | Not a health facility | Not a health facility | | other private medical sector |
| | | | | pharmacy |
| | | | | shop |
| | | | | firend/relative |
| | | | | other |
| | | | | don't know |
| Tanzania | | | SPA 2014-15 | DHS 2015-16 |
| | Public/Parastatal | Government/public/parastatal hopistal | national referral hospital | national/zonal referral/spec. hospital (government/parastatal) |
| | | | regional hospital | regional referral hospital (government/parastatal) |
| | | | district hospital | regional hospital (government/parastatal) |
| | | | district-designated hospital | district hospital (government/parastatal) |
| | | | other hospital (private) | |
| | | Government/public/parastatal health center | health centre | health center (government/parastatal) |
| | | Government/public/parastatal health clinic/dispensary | clinic | dispensary (government/parastatal) |
| | | | dispensary | clinic (government/parastatal) |
| | | | | chw (government/parastatal) |
| | Private | Private for profit | other hospital (private) | specialized hospital (private) |
| | | | health centre | hospital (private) |
| | | | clinic | health centre (priavte) |
| | | | dispensary | dispensary (priavte) |
| | | | | clinic (private) |
| | | Mission/faith-based | national referral hospital | referal/sepc. hospital (religious/voluntary) |
| | | | regional hospital | district hospital (religious/voluntary) |
| | | | district hospital | hospital (religious/voluntary) |
| | | | district-designated hospital | health centre (religious/voluntary) |
| | | | other hospital (private) | dispensary (religious/voluntary) |
| | | | health centre | clinic (religious/voluntary) |
| | | | clinic | ngo |
| | | | dispensary | |
| | Not a health faciliy | Not a health faciliy | | pharmacy |
| | | | | acredited drug dispensing outlet (addo) |
| | | | | shop/kiosk |
| | | | | bar |
| | | | | guest house/hotel |
| | | | | friend/relative/neighor |
| | | | | vct centre |
| | | | | other |
