## Appendix 2 for "Inequities in Effective Coverage of Family Planning Services in Low-and Middle-Income Countries: Linking Households and Facility Surveys"

Appendix 2. Formal Source for Modern Method

| Country | Geographic area, Facility type | Proportion | Sample Size |
|---|---|---|---|
| **Bangladesh** | urban, public hosp | 1% | 111 |
| | urban, public UHC | 4% | 251 |
| | urban, public MCWC | 1% | 107 |
| | urban, public UnHFWC | 1% | 76 |
| | urban, public clinic/post | 9% | 587 |
| | urban, private for profit | 3% | 182 |
| | urban, private for non-profit | 4% | 355 |
| | rural, public hosp | 2% | 120 |
| | rural, public UHC | 12% | 559 |
| | rural, public MCWC | 2% | 108 |
| | rural, public UnHFWC | 7% | 345 |
| | rural, public clinic/post | 43% | 2010 |
| | rural, private for profit | 6% | 262 |
| | rural, private for non-profit | 4% | 203 |
| **Haiti** | Urban, public hosp | 12% | 254 |
| | Urban, public HC | 16% | 315 |
| | Urban, public clinic/post | 1% | 20 |
| | Urban, private for profit | 6% | 111 |
| | Urban, private for non-profit | 5% | 122 |
| | Rural, public hosp | 14% | 353 |
| | Rural, public HC | 26% | 709 |
| | Rural, public clinic/post | 1% | 15 |
| | Rural, private for profit | 5% | 133 |
| | Rural, private for non-profit | 15% | 417 |
| **Malawi** | North, public hosp | 2% | 561 |
| | North, public HC | 6% | 768 |
| | North, public clinic/post | 2% | 250 |
| | North, private for profit | 0% | 29 |
| | North, private for non-profit | 1% | 440 |
| | Central, public hosp | 7% | 822 |

| | | | |
|---|---|---|---|
| | Central, public HC | 24% | 1,870 |
| | Central, public clinic/post | 6% | 468 |
| | Central, private for profit | 3% | 218 |
| | Central, private for non-profit | 5% | 435 |
| | South, public hosp | 6% | 999 |
| | South, public HC | 23% | 2,480 |
| | South, public clinic/post | 5% | 512 |
| | South, private for profit | 3% | 238 |
| | South, private for non-profit | 6% | 600 |
| **Nepal 2016** | Mountain, public hosp | 2% | 80 |
| | Mountain, public HC | 0% | 19 |
| | Mountain, public post | 4% | 168 |
| | Mountain, private for profit | 0% | 7 |
| | Mountain, private for non-profit | 0% | 3 |
| | Hill, public hosp | 12% | 483 |
| | Hill, public HC | 1% | 53 |
| | Hill, public post | 19% | 802 |
| | Hill, private for profit | 6% | 188 |
| | Hill, private for non-profit | 3% | 78 |
| | Terai, public hosp | 23% | 794 |
| | Terai, public HC | 2% | 84 |
| | Terai, public post | 16% | 625 |
| | Terai, private for profit | 6% | 204 |
| | Terai, private for non-profit | 4% | 137 |
| **Nepal 2022** | Mountain, public hosp | 2% | 139 |
| | Mountain, public HC | 1% | 54 |
| | Mountain, public post | 4% | 258 |
| | Mountain, private for profit | 0% | 14 |
| | Mountain, private for non-profit | 0% | 7 |
| | Hill, public hosp | 10% | 465 |
| | Hill, public HC | 2% | 103 |
| | Hill, public post | 17% | 965 |
| | Hill, private for profit | 5% | 170 |

| | | | |
|---|---|---|---|
| | Hill, private for non-profit | 1% | 69 |
| | Terai, public hosp | 20% | 713 |
| | Terai, public HC | 1% | 36 |
| | Terai, public post | 24% | 942 |
| | Terai, private for profit | 9% | 315 |
| | Terai, private for non-profit | 4% | 142 |
| **Tanzania** | Mainland urban, public/parastatal hosp | 9% | 194 |
| | Mainland urban, public/parastatal HC | 8% | 171 |
| | Mainland urban, public/parastatal clinic/dispensary | 8% | 186 |
| | Mainland urban, private for profit | 4% | 67 |
| | Mainland urban, mission/faith-based | 3% | 72 |
| | Mainland rural, public/parastatal hosp | 6% | 147 |
| | Mainland rural, public/parastatal HC | 11% | 223 |
| | Mainland rural, public/parastatal clinic/dispensary | 42% | 998 |
| | Mainland rural, private for profit | 2% | 41 |
| | Mainland rural, mission/faith-based | 8% | 162 |

Appendix 3. Readiness Score by Linking Unit

| Country | Geographic area, Facility type | Readiness Score | Sample Size |
|---|---|---|---|
| Bangladesh | Urban, public hosp | 0.74 | 51 |
| | Urban, public UHC | 0.94 | 72 |
| | Urban, public MCWC | 0.90 | 71 |
| | Urban, public UnHFWC | 0.77 | 5 |
| | Urban, public clinic/post | 0 | 0 |
| | Urban, private for profit | 0.49 | 57 |
| | Urban, private for non-profit | 0.82 | 68 |
| | Rural, public hosp | 0 | 0 |
| | Rural, public UHC | 0.89 | 62 |
| | Rural, public MCWC | 0.82 | 19 |
| | Rural, public UnHFWC | 0.76 | 600 |
| | Rural, public clinic/post | 0.50 | 302 |
| | Rural, private for profit | 0.58 | 4 |
| | Rural, private for non-profit | 0.78 | 42 |
| Haiti | Urban, public hosp | 0.86 | 45 |
| | Urban, public HC | 0.71 | 76 |
| | Urban, public clinic/post | 0.78 | 15 |
| | Urban, private for profit | 0.58 | 75 |
| | Urban, private for non-profit | 0.66 | 33 |
| | Rural, public hosp | 0.79 | 10 |
| | Rural, public HC | 0.68 | 119 |
| | Rural, public clinic/post | 0.64 | 211 |
| | Rural, private for profit | 0.58 | 111 |
| | Rural, private for non-profit | 0.59 | 61 |
| Malawi | North, public hosp | 0.85 | 13 |
| | North, public HC | 0.77 | 73 |
| | North, public clinic/post | 0.52 | 11 |
| | North, private for profit | 0.54 | 20 |
| | North, private for non-profit | 0.72 | 25 |
| | Central, public hosp | 0.81 | 16 |

| | | | |
|---|---|---|---|
| | Central, public HC | 0.69 | 131 |
| | Central, public clinic/post | 0.52 | 22 |
| | Central, private for profit | 0.58 | 84 |
| | Central, private for non-profit | 0.63 | 57 |
| | South, public hosp | 0.85 | 20 |
| | South, public HC | 0.72 | 139 |
| | South, public clinic/post | 0.46 | 32 |
| | South, private for profit | 0.57 | 108 |
| | South, private for non-profit | 0.70 | 59 |
| **Nepal 2015** | Mountain, public hosp | 0.91 | 17 |
| | Mountain, public HC | 0.78 | 23 |
| | Mountain, public post | 0.57 | 86 |
| | Mountain, private for profit | 0.48 | 5 |
| | Mountain, private for non-profit | 0.63 | 4 |
| | Hill, public hosp | 0.85 | 51 |
| | Hill, public HC | 0.79 | 117 |
| | Hill, public post | 0.62 | 192 |
| | Hill, private for profit | 0.64 | 45 |
| | Hill, private for non-profit | 0.70 | 22 |
| | Terai, public hosp | 0.82 | 33 |
| | Terai, public HC | 0.75 | 105 |
| | Terai, public post | 0.61 | 145 |
| | Terai, private for profit | 0.57 | 35 |
| | Terai, private for non-profit | 0.78 | 19 |
| **Nepal 2021** | Mountain, public hosp | 0.82 | 21 |
| | Mountain, public HC | 0.50 | 102 |
| | Mountain, public post | 0.68 | 62 |
| | Mountain, private for profit | 0.44 | 7 |
| | Mountain, private for non-profit | 0.36 | 1 |
| | Hill, public hosp | 0.84 | 71 |
| | Hill, public HC | 0.52 | 416 |
| | Hill, public post | 0.62 | 209 |
| | Hill, private for profit | 0.57 | 83 |

| | | | |
|---|---|---|---|
| | Hill, private for non-profit | 0.70 | 23 |
| | Terai, public hosp | 0.85 | 45 |
| | Terai, public HC | 0.56 | 235 |
| | Terai, public post | 0.63 | 109 |
| | Terai, private for profit | 0.57 | 73 |
| | Terai, private for non-profit | 0.78 | 21 |
| **Tanzania** | Mainland urban, public/parastatal hosp | 0.91 | 92 |
| | Mainland urban, public/parastatal HC | 0.85 | 56 |
| | Mainland urban, public/parastatal clinic/dispensary | 0.66 | 30 |
| | Mainland urban, private for profit | 0.74 | 54 |
| | Mainland urban, mission/faith-based | 0.74 | 35 |
| | Mainland rural, public/parastatal hosp | 0.87 | 19 |
| | Mainland rural, public/parastatal HC | 0.79 | 197 |
| | Mainland rural, public/parastatal clinic/dispensary | 0.62 | 298 |
| | Mainland rural, private for profit | 0.65 | 17 |
| | Mainland rural, mission/faith-based | 0.78 | 56 |

Appendix 4. Informal Source for Modern Method by Geographic Area, Women's age, and Household Wealth

| **Country** | | **Sample size** | **Proportion** |
|---|---|---|---|
| **Bangladesh** | Urban | 2097 | 56% |
| | Rural | 2505 | 41% |
| | 15-17 | 188 | 71% |
| | 18-24 | 1455 | 62% |
| | 25-34 | 1930 | 47% |
| | 35-49 | 1029 | 32% |
| | Poorest | 718 | 36% |
| | Richest | 1365 | 65% |
| **Haiti** | Urban | 487 | 37% |
| | Rural | 215 | 12% |
| | 15-17 | 72 | 76% |
| | 18-24 | 308 | 37% |
| | 25-34 | 223 | 18% |
| | 35-49 | 99 | 10% |
| | Poorest | 37 | 6% |
| | Richest | 267 | 48% |
| **Malawi** | Northern | 38 | 2% |
| | Central | 101 | 3% |
| | Southern | 141 | 3% |
| | 15-17 | 49 | 20% |
| | 18-24 | 133 | 5% |
| | 25-34 | 64 | 1% |
| | 35-49 | 34 | 1% |
| | Poorest | 28 | 2% |
| | Richest | 136 | 5% |
| **Nepal 2015** | Mountain | 21 | 7% |

| | | | |
|---|---|---|---|
| | Hill | 237 | 13% |
| | Terai | 322 | 15% |
| | 15-17 | 8 | 29% |
| | 18-24 | 102 | 20% |
| | 25-34 | 241 | 15% |
| | 35-49 | 229 | 10% |
| | Poorest | 68 | 8% |
| | Richest | 186 | 26% |
| **Nepal 2021** | Mountain | 25 | 5% |
| | Hill | 300 | 14% |
| | Terai | 276 | 11% |
| | 15-17 | 9 | 35% |
| | 18-24 | 104 | 19% |
| | 25-34 | 234 | 13% |
| | 35-49 | 254 | 10% |
| | Poorest | 107 | 8% |
| | Richest | 176 | 29% |
| **Tanzania** | Mainland Urban | 396 | 36% |
| | Mainland Rural | 334 | 18% |
| | 15-17 | 41 | 58% |
| | 18-24 | 231 | 31% |
| | 25-34 | 288 | 26% |
| | 35-49 | 170 | 16% |
| | Poorest | 42 | 11% |
| | Richest | 311 | 42% |
